# Relationship between finger movement characteristics and voxel-based specific regional analysis systems for Alzheimer’s disease

**DOI:** 10.1101/2022.05.23.22275382

**Authors:** Junpei Sugioka, Shota Suzumura, Katsumi Kuno, Shiori Kizuka, Hiroaki Sakurai, Yoshikiyo Kanada, Tomohiko Mizuguchi, Izumi Kondo

## Abstract

**Background:** Aging is the most significant risk factor of dementia. Alzheimer’s disease accounts for approximately 60%–80% of all dementia cases in elderly individuals. This study aimed to examine the relationship between finger movements and brain function in patients with Alzheimer’s disease, using VSRAD, and explore the relationship between VSRAD and cognitive function (MoCA-J).

**Methods:** Patients diagnosed with AD, at the Center for Comprehensive Care and Research on Memory Disorders, were included in the study. The diagnostic criteria were based on the National Institute on Aging-Alzheimer’s Association. Patients were excluded based on set criteria. A finger-tapping device was used for all measurements. Participants performed the following task, in order: non-dominant hand, dominant hand, simultaneously, and alternate hands. Movements were measured for 15 s each. The relationship between distance and output was measured. MRI measurements were performed, after which VSRAD was performed using sagittal section 3D T1-weighted images. The Z-score was used to calculate the degree of atrophy. Pearson’s product-moment correlation coefficient was used to analyze the relationship between the Z-score and mean values of the parameters in the finger-tapping movements. Statistical significance was set at p < 0.05.

**Results:** Sixty-two patients were included in the study. Z-scores for MoCA-J analysis, percentage of total brain atrophy in the white matter, and other VSRAD parameters showed varying negative correlations (r = -0. 28, p = 0. 035, r = -0. 31, p = 0. 020, and r = -0. 52, p < 0. 001), respectively. Varying positive correlations between Z-score and the SD of distance rate of velocity peak in extending movements in the non-dominant hand were observed.

**Conclusion:** The SD of distance rate of velocity peak in extending movements and the Z-score showed a significant relationship. An association between neuropsychological tests and overall degree of brain atrophy was also observed.

## Introduction

Aging is the most significant risk factor of dementia. Since the latter half of the 20th century, amidst global aging, the average life expectancy of Japanese men and women has reached 84.5 years and 90.6 years, respectively [1]. In the future, the number of people with dementia is expected to increase as the population continues to age. It is difficult to cure dementia using current medical science; however, early diagnosis and intervention can potentially prevent dementia [2]. Alzheimer’s disease (AD) accounts for approximately 60%–80% of all cases of dementia in elderly individuals [3]. It is also reported that approximately 50% of patients with mild cognitive impairment (MCI), which falls between dementia and being healthy, will progress to AD within five years [4]. Therefore, the early detection of AD is important to prevent the onset and progression of dementia.

Against this social background, several studies have been conducted on the early diagnosis of AD, and it has become possible to make an early diagnosis of dementia to some extent. Diagnosing dementia includes imaging tests, such as magnetic resonance imaging (MRI) [5], single-photon emission computed tomography [6], fluorodeoxyglucose-positron emission tomography [7], and cerebrospinal fluid biomarkers [8]. Particularly, voxel-based specific regional analysis systems for Alzheimer’s disease (VSRAD) have recently gained attention as tools for the early diagnosis of AD. VSRAD are a software program that automatically calculates the degree of atrophy in the medial temporal and dorsal brainstem from MRI images [9]. A study using VSRAD reported that the diagnostic accuracy of AD improved by using VSRAD and the Mini-Mental State Examination (MMSE) for combined diagnosis [10]. Additionally, an observational study of community-dwelling elderly individuals reported that the degree of medial temporal atrophy (Z-score) is related to the amount of activity [11]. The Montreal Cognitive Assessment (MoCA) is becoming increasingly popular in clinical practice as a superior screening test for the detection of mild AD and MCI. Although MoCA is said to be more sensitive and specific than MMSE for detecting patients with mild AD or MCI [12], there are still no reports on the association between VSRAD and MoCA.

However, it has recently been reported that motor impairment can be detected in the early stages of dementia and MCI and that there is a possibility that signs of dementia can be detected from motor impairment. Verghese, Robbins, & Holtzer et al. (2008) conducted a quantitative gait evaluation and reported a decrease in walking ability, including walking speed and stride length, in patients with MCI compared to healthy elderly people [13]. We have been conducting preliminary studies on finger movements in patients with dementia based on the fact that hand movements may be able to detect pathological changes in the brain at an early stage. Accordingly, we detected finger movement features that appear with cognitive decline and that finger dexterity declines in the stages of AD and MCI compared to healthy conditions [14-17]. Additionally, in a study of cognitive function and hand function at other institutions, it was reported that the number of finger tappings decreased and that the tapping interval was longer in patients with AD and those with MCI than in healthy elderly subjects [18]. The number of studies examining hand function in patients with AD or MCI, including our study, is increasing. However, no reports have examined the relationship between finger function and brain imaging in patients with dementia. It is assumed that finger movements are intricately related to various parts of the brain and that there are many aspects that have not been investigated yet.

Thus, this study aimed to examine the relationship between finger movements and brain function in patients with AD using VSRAD and explore the relationship between VSRAD and cognitive function.

## Methods

### Research Design and Subjects

This exploratory, cross-sectional study was conducted at the National Center for Geriatrics and Gerontology. This study included patients diagnosed with AD at the Center for Comprehensive Care and Research on Memory Disorders. The diagnostic criteria for AD were based on the National Institute on Aging-Alzheimer’s Association [19]. If a definite diagnosis could not be reached, the diagnosis was discussed at a conference of specialists in dementia and subsequently made. The exclusion criteria were impaired consciousness, tremor, parkinsonism, higher brain dysfunction, such as aphasia or apraxia, epilepsy, paralysis, sensory disturbance, finger dexterity impairment, severe cognitive dysfunction that made neuropsychological testing difficult, left-handedness, and difficulty in MRI measurements.

### Ethical consideration

This study was conducted with the approval of the Ethics and Conflict of Interest Committee of the National Center for Geriatrics and Gerontology (approval number: 1485-2). The purpose of this study was explained in advance to the subjects, orally and in writing, and only those who provided consent were included in the study.

### Finger tapping measurement and cognitive function assessment

Finger function was measured as finger-tapping movements. Finger-tapping movements were defined as repetitive opening and closing movements by the thumb and index finger. We used a finger-tapping device with magnetic sensors (UB-2, Maxell Holdings, Ltd., Tokyo, Japan) for measurement (Fig 1). Magnetic sensors were attached to the dorsal surface of the thumb and index finger, and finger tapping movements were measured. Measurements were performed in the following order: non-dominant hand, dominant hand, simultaneously, and alternate hands. Movements were measured for 15 s (total time for all four movements, 60 s) (Fig 2). The magnetic sensor finger tapping device can calculate 44 parameters after measuring and recording the finger tapping motion at 0.1-s intervals (Table 1). The relationship between distance and output was measured as the output characteristics of the device. The slope of the regression line and R^2^ value were 0.9991 and 0.9999, respectively (Fig 3).

**Table 1.**
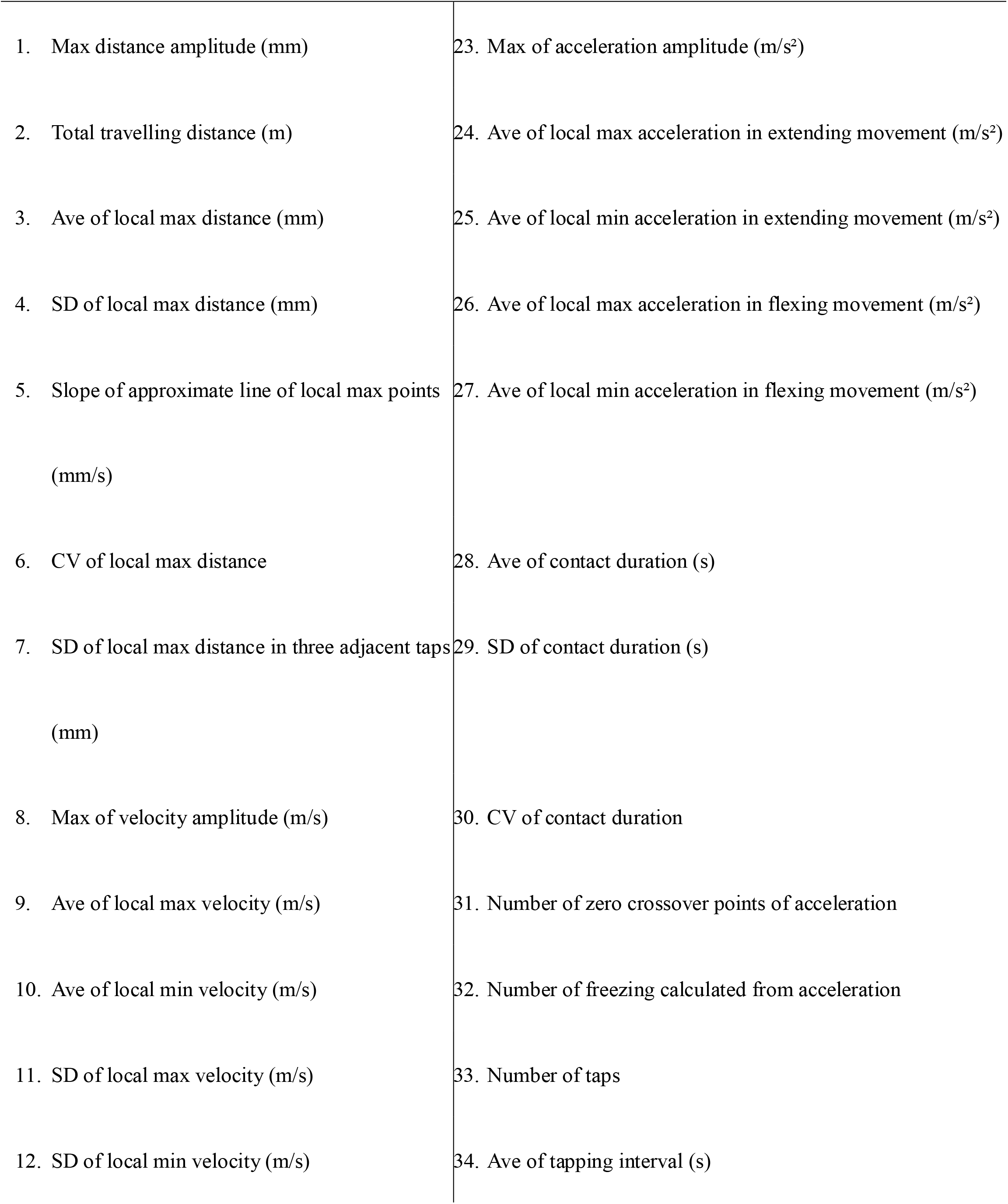
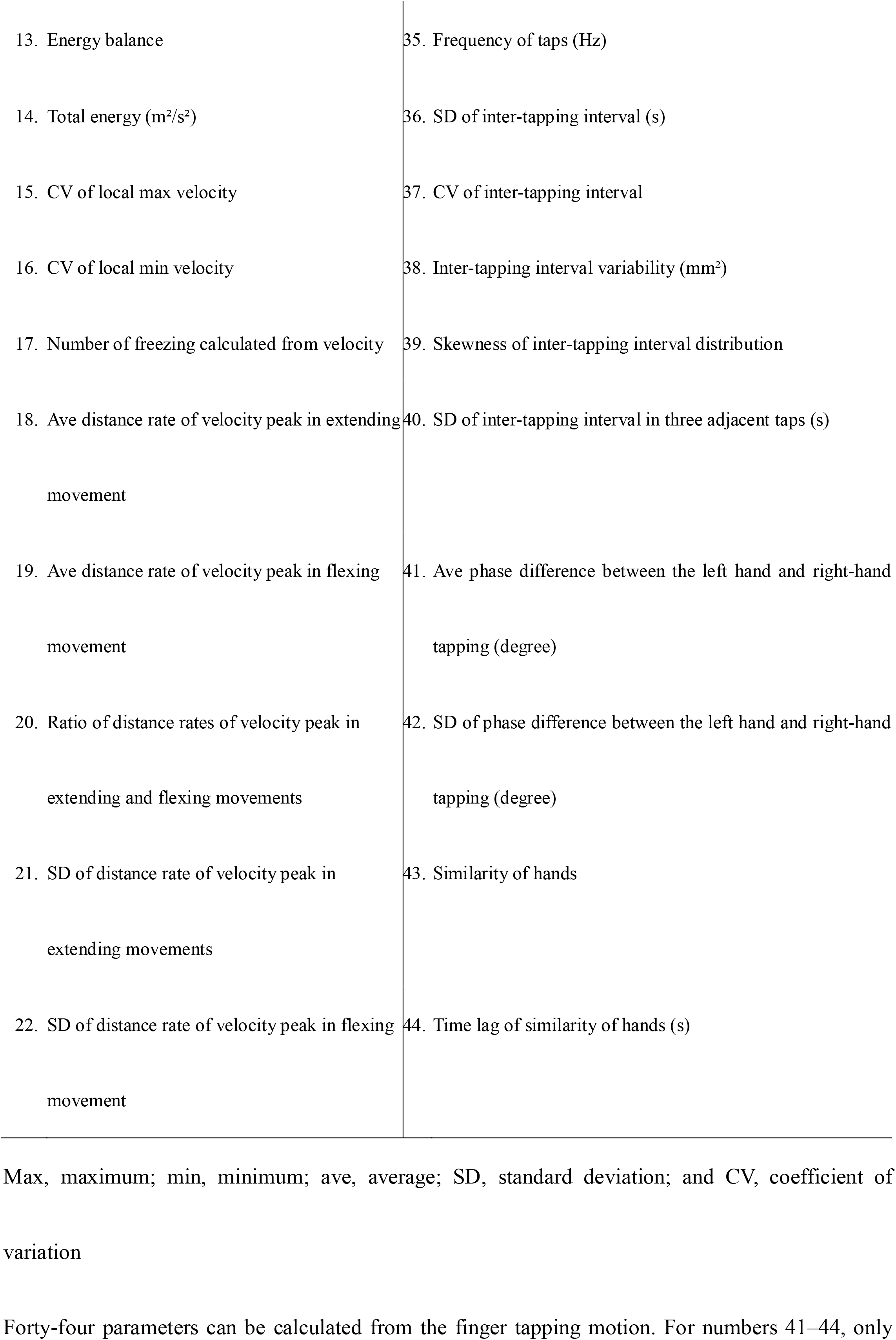
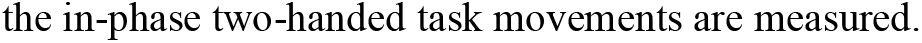
Parameter of finger tapping movements.

**Fig 1.**
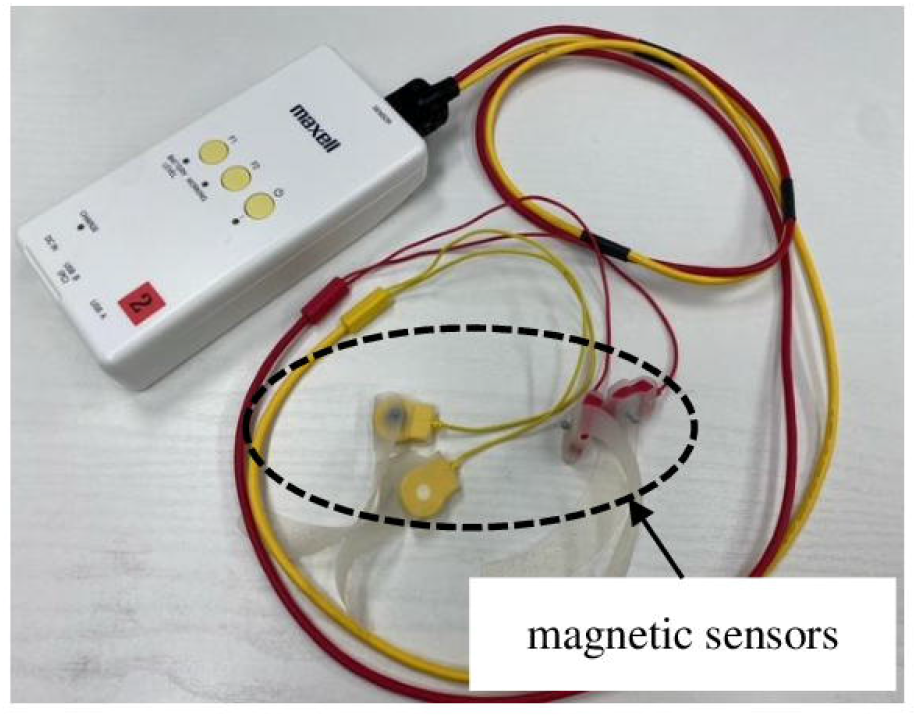
The UB-2 finger tapping device with magnetic sensors. Yellow cables were used for the left-hand side, and red cables were used for the right-hand side. The magnetic sensor was attached to the dorsal surface of the thumb and index finger and fixed with a rubber belt.

**Fig 2.**
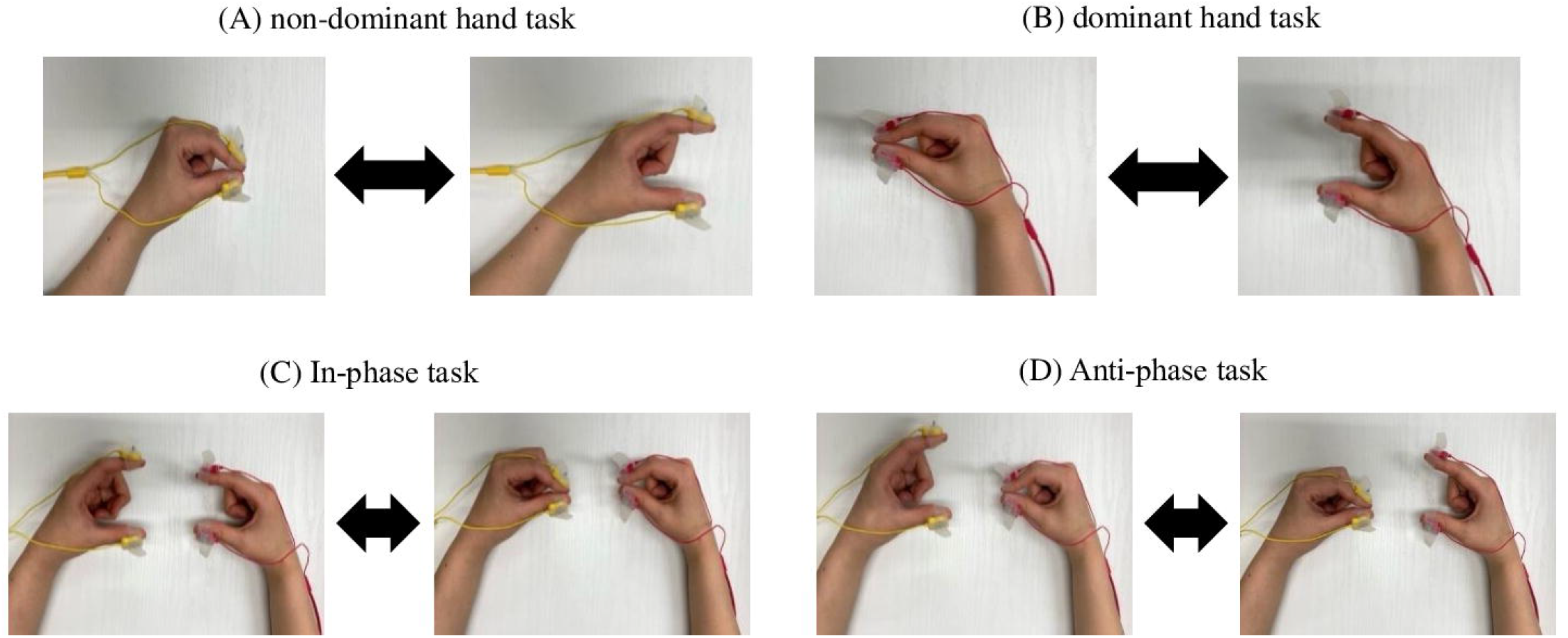
Finger tapping movements. (A) A tapping action with the left thumb and index finger is perfomed. (B) A tapping action with the right thumb and index finger is performed. (C) Tapping with the in-phase task is performed. (D) Tapping with the anti-phase task is performed. The distance between the thumb and index finger is kept between 3 and 4 cm, and movement was performed as fast as possible for 15 s.

**Fig 3.**
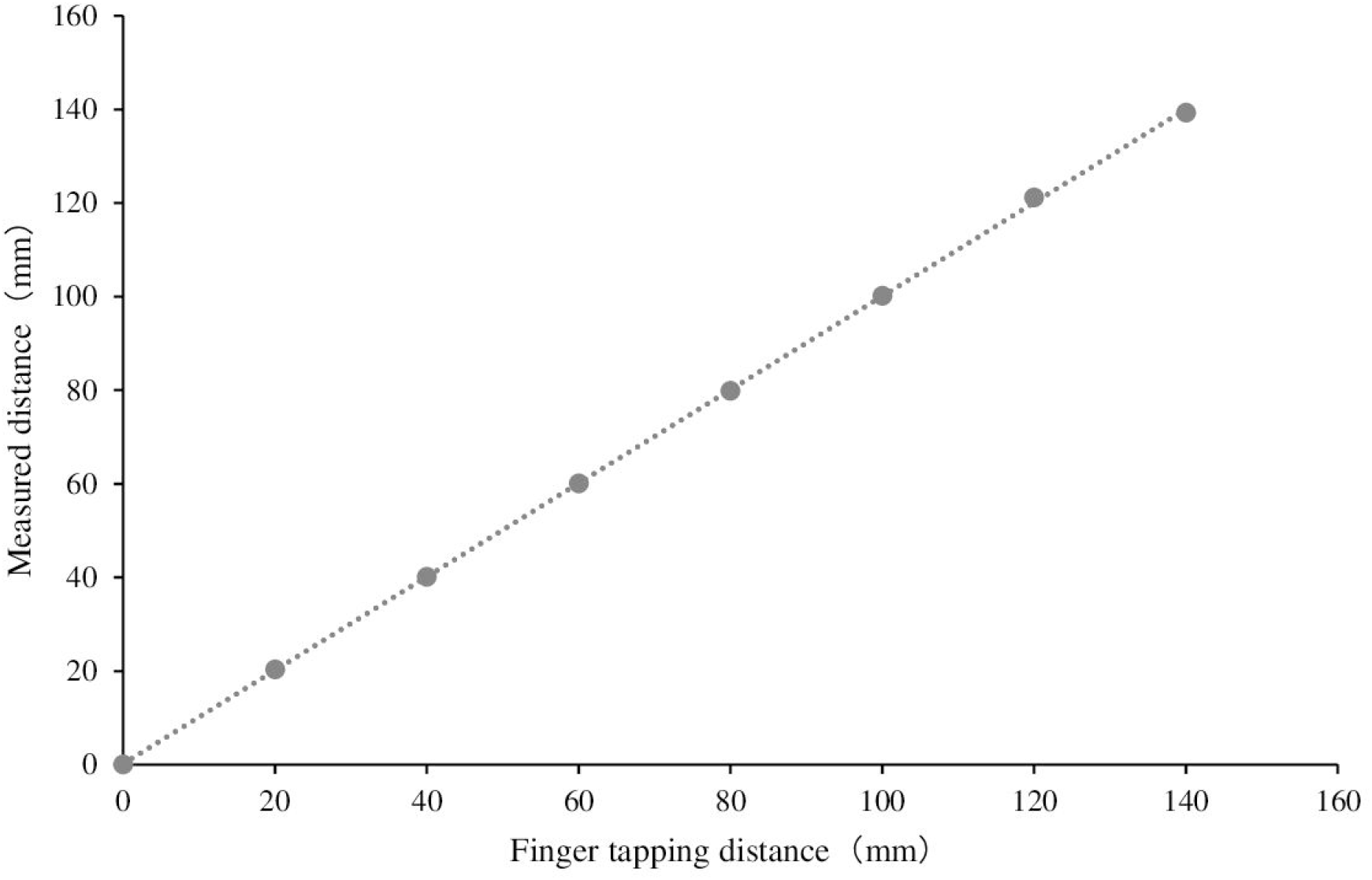
Output characteristics of the measuring equipment. Figure 3 shows the relationship between the finger tapping distance (mm) and measured distance (mm) of the device. The slope of the regression line and the R^2^ value were 0.9991 and 0.9999, respectively.

Additionally, the Japanese version of the MoCA (MoCA-J) [12], which can quantitatively evaluate the severity of cognitive function in general, was given to all participants.

### Brain MRI measurement and data acquisition

MRI measurements were performed using an Ingenia Ambition 1.5T scanner (Philips Japan, Tokyo). After MRI measurements, a VSRAD analysis (VSRAD advance 2, Eisai Co., Ltd, Tokyo, Japan) was performed using sagittal section 3D T1-weighted images. VSRAD is a software program that measures brain atrophy by volume and automatically calculates the degree of atrophy in the medial temporal region and dorsal brainstem by computing image information obtained by MRI [9]. VSRAD can be measured simply by adding the imaging conditions for analysis to the normal MRI imaging and can be performed in about 20–30 minutes with little burden on the human body and no additional charges on insurance. The indices were divided into gray and white matter. The gray matter indices were calculated as follows: (1) degree of atrophy within the volume of interest (VOI), (2) percentage of total brain atrophy, (3) percentage of atrophy within VOI, (4) atrophy ratio (VOI/total brain atrophy percentage), and (5) maximum value within VOI. The white matter indices were calculated as the percentage of total brain atrophy (Table 2). The region of interest to be used as a reference to support the assessment of brain atrophy in patients with AD was the medial temporal cortex (hippocampus, amygdala, and most of the olfactory cortex). The degree of atrophy within the VOI was indicated by the Z-score. The Z-score indicates how much the standard deviation (SD) is separated from the mean value by statistically comparing the subject’s brain image and a normal brain image. As a guide, a Z-score of 0 to 1 shows almost no atrophy in the region of interest, a Z-score 1 to 2 shows some atrophy in the region of interest, and a Z-score of 2 to 3 shows considerable atrophy in the region of interest. If the value is larger than 3, it is judged that the atrophy in the region of interest is strong. The results of the VSRAD analysis were obtained from MRI data performed in an outpatient memory clinic.

**Table 2.** Results calculated by VSRAD.

| Item | Parameter | Total value | Right side<br>value | Left side<br>value | Right side -<br>Left side |
| --- | --- | --- | --- | --- | --- |
| Gray matter ① | Severity of VOI atrophy | ○ | ○ | ○ | ○ |
| Gray matter ② | Extent of GM atrophy (%) | ○ | — | — | — |
| Gray matter ③ | Extent of VOI atrophy (%) | ○ | ○ | ○ | ○ |
| Gray matter ④ | Ratio of VOI/GM atrophy (times) | ○ | — | — | — |
| Gray matter ⑤ | Max in VOI | ○ | ○ | ○ | ○ |
| White matter | Extent of WM atrophy (%) | ○ | — | — | — |

○, calculable; —, not calculated
Gray matter ① Average of Z-scores above 0 in VOI
Gray matter ② Percentage of areas with a Z-score >2 within the total gray matter
Gray matter ③ Percentage of regions in a VOI having a Z-score of >2
Gray matter ④ Percentage of cases with a total brain atrophy value of 1
White matter Percentage of areas with a Z-score >2 within the total white matter

### Statistical analysis

The association between the values calculated by the VSRAD analysis and MOCA-J was analyzed using Spearman’s rank correlation coefficient. After the finger-tapping movements were measured, the measured values of all 44 parameters were calculated from the just tap. Pearson’s product-moment correlation coefficient was used to analyze the relationship between the Z-score and mean values of the parameters in the finger-tapping movements. A p-value <0.05 was considered statistically significant. Statistical analysis was performed using SPSS Statistics version 26.0 (IBM Corp., Armonk, NY, USA).

## Results

### Participant characteristics

Measurements were performed on 68 individuals. Six left-handed participants were excluded owing to the exclusion criteria, and 62 participants were analyzed. Additionally, five patients were unable to perform the MoCA-J test due to difficulty in understanding the instructions. Thus, 57 patients were included in the MoAC-J analysis (Table 3).

**Table 3.**
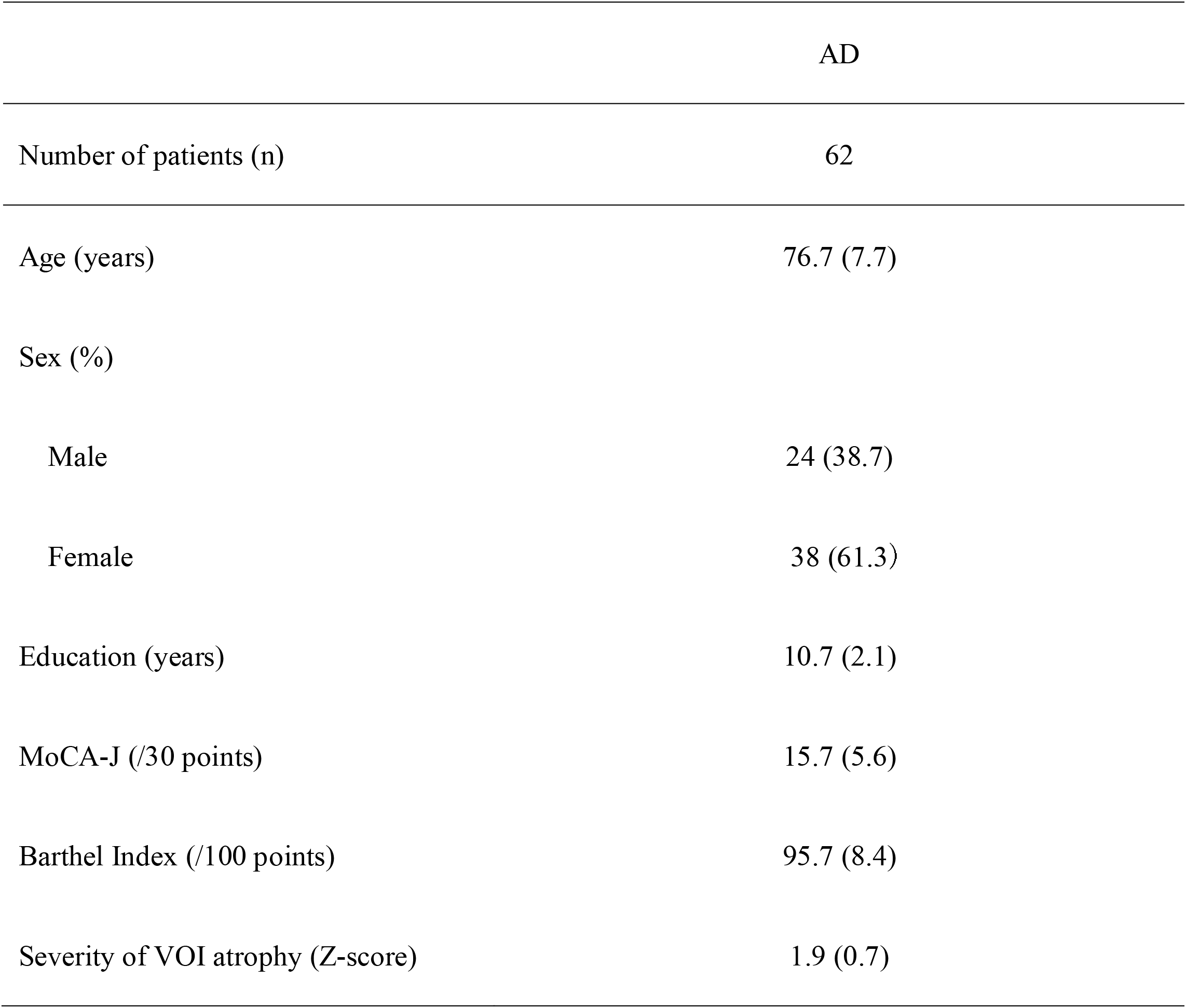
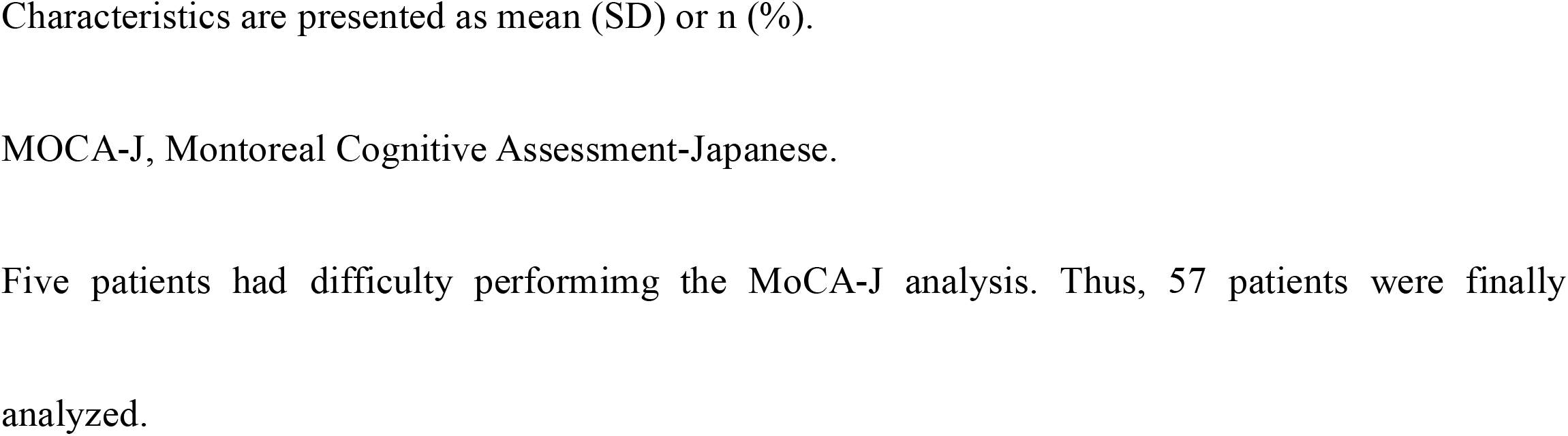
Patient Characteristics.

### Relationship between VSRAD and MoCA-J

The MoCA-J analysis showed a significant weak negative correlation with the Z-score (r=-0.28 and p=0.035). The other VSRAD parameters showed a significant negative correlation with the percentage of total brain atrophy in the gray matter (r=-0. 52 and p<0.001). A significant weak negative correlation was observed between the percentage of total brain atrophy in the white matter (r=-0. 31 and p=0. 020) (Fig 4).

**Fig 4.**
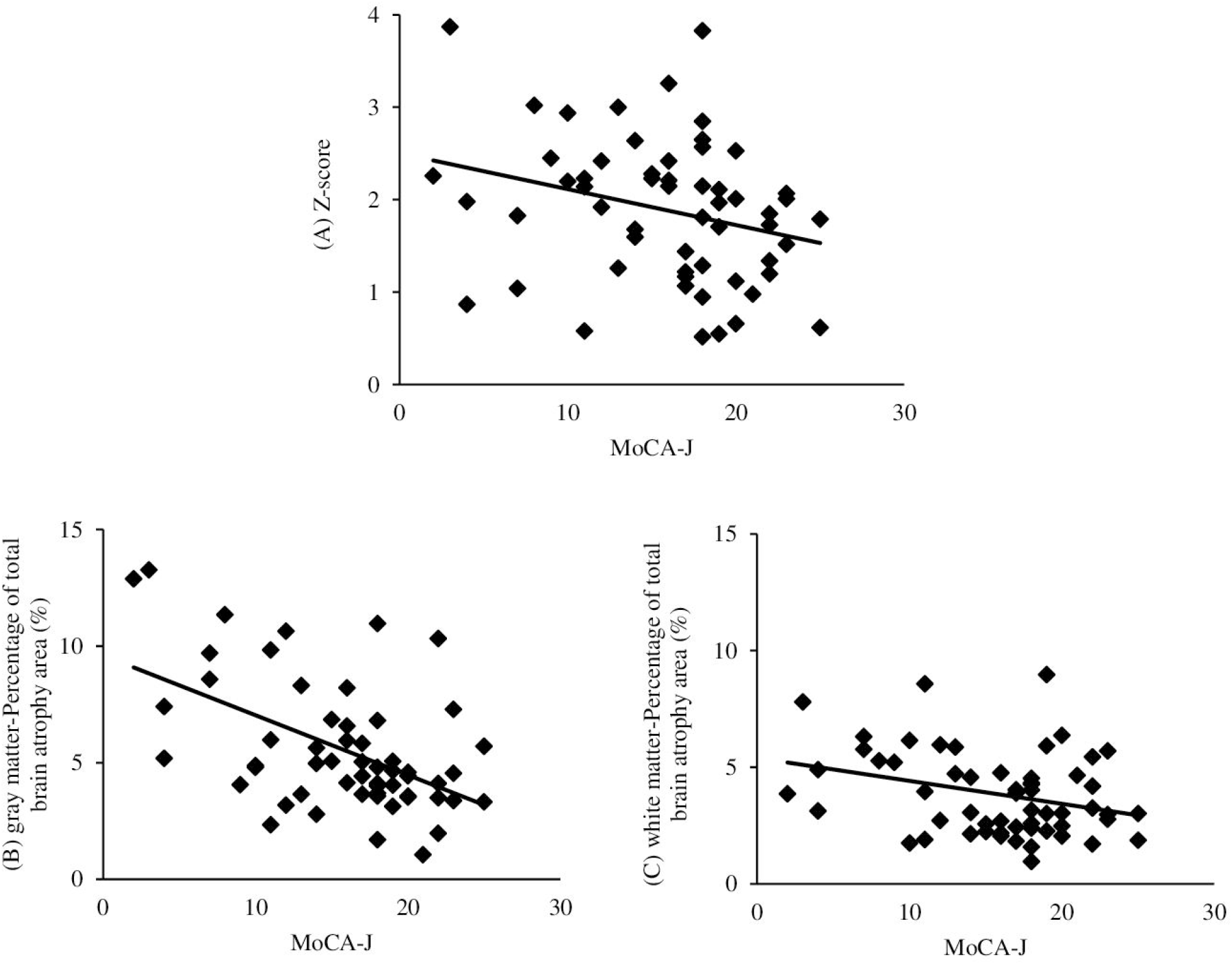
Relationship between the VSRAD analysis index and MoCA-J. The vertical axis indicates the VSRAD analysis index, and the horizontal axis indicates the MoCA-J scores. (A) Zscore: r=-0. 28 and p=0. 035. (B) Percentage of total brain atrophy in the gray matter: r=-0. 52 and p <0. 001. (C) Percentage of total brain atrophy in the white matter: r=-0. 31 and p=0.020.

### Relationship between VOI internal atrophy (Z-score) and finger-tapping movements

A positive correlation was recognized between the Z-score and SD of the distance rate of velocity peak in extending movements in the non-dominant hand (r=0. 51 and p<0.001). Moreover, we recognized weak positive correlations between the SD of the local max distance in non-dominant movements (r=0. 26 and p=0. 035) and the SD of contact duration of the anti-phase task in the non-dominant hand (r=0. 25 and p=0. 044) (Fig 5). No significant correlation was found between other finger tap parameters and the Z-score.

**Fig 5.**
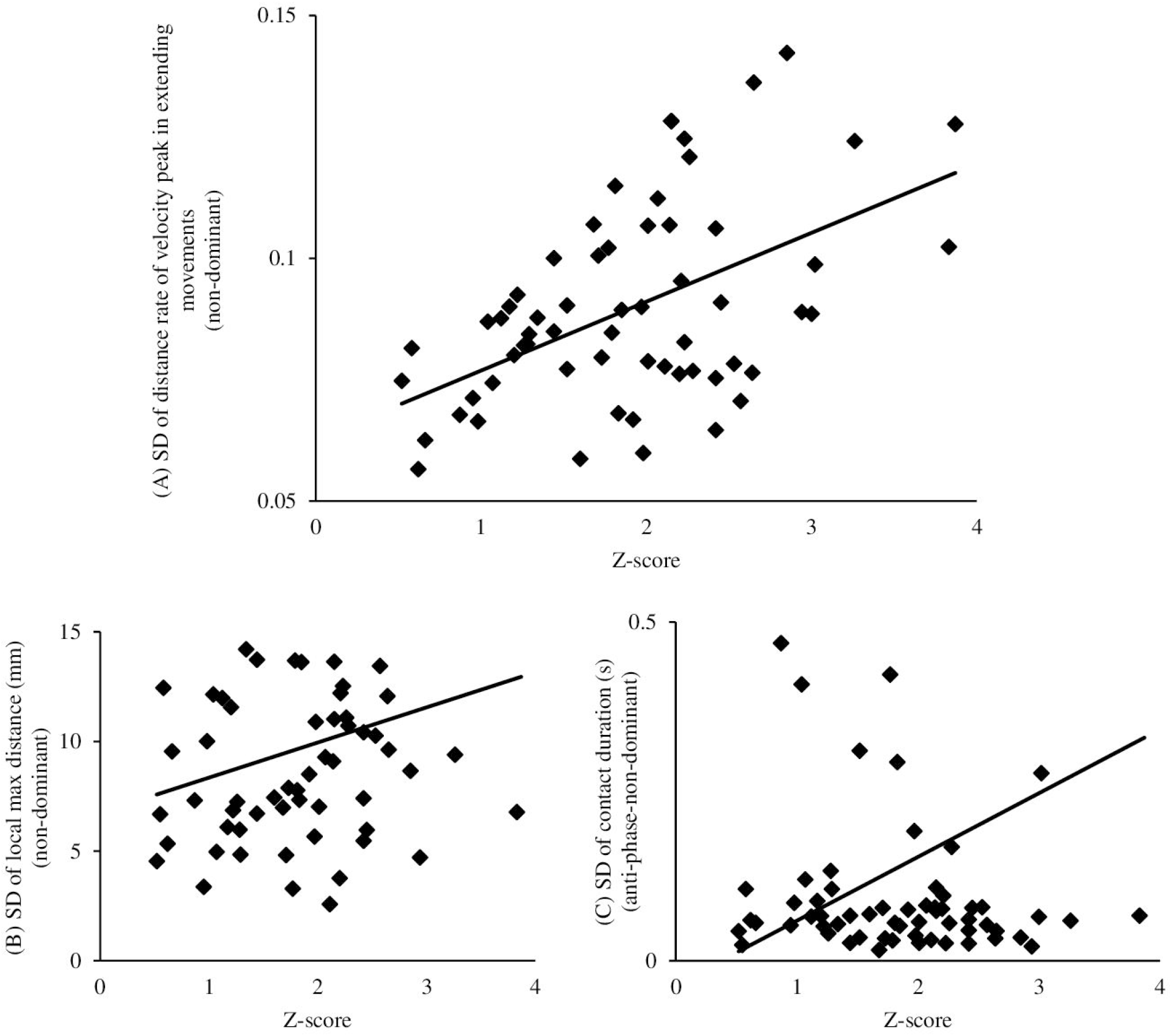
Relationship between finger-tapping parameters and Z-score. The vertical and horizontal axes indicate the parameters of the finger tapping movements and Z-score values, respectively. (A) r=0. 51 and p <0. 001. (B) r=0. 26 and p=0. 035. (C) r=0.25 and p=0.044.

The SD of distance rate of velocity peak in extending movements is defined as the SD of the value calculated as the position of the distance at which the velocity is maximal during the finger opening movement as a ratio to the amplitude.

The SD of local max distance is defined as the SD of the amplitude (maximum value per one-time tap) of the distance waveform.

The SD of contact duration is defined as the standard deviation of the time at which the index finger and thumb are closed.

## Discussion

We examined the relationship between finger function, brain function, and cognitive function in patients with AD using VSRAD. The results showed a significant correlation between the SD of distance rate of velocity peak in extending movements and the Z-score (r=0. 51 and p <0. 001). A weak correlation was observed between the SD of local max distance (r=0. 26 and p=0. 035) and the SD of contact duration (r=0. 25 and p=0. 044). The MoCA-J analysis showed significant correlations between the Z-score (r=-0. 28 and p=0. 035), percentage of gray matter atrophy (r=-0. 52 and p <0. 001), and percentage of white matter atrophy (r=-0. 31 and p=0. 020).

Although finger movements have been reported to be associated with various brain areas and are still unclear in many cases, to the best of our knowledge, no reports have verified the relationship between finger function and brain imaging analysis in patients with dementia. Although the Brodmann area is well known for its functional localization in the brain, the areas that control finger functions are related to the functions of the primary motor cortex (BA4), premotor cortex, and supplementary motor area (BA6). In addition to these areas, other brain areas, such as the primary somatosensory cortex (BA3, BA1, and BA2) and cerebellum, are involved in a complex manner in finger movement [20-22]. Conversely, the hippocampus, amygdala, and entorhinal cortex participate in motor speed and acceleration during visuomotor tasks, in addition to memory and emotion [23, 24]. Areas other than the motor cortex, such as the medial temporal region, are also involved in movement; Therefore, brain atrophy due to AD [25] may affect finger movements.

In this study, a significant correlation was found between the SD of distance rate of velocity peak in extending movements and Z-scores. This parameter is the variation in the point of maximum velocity while opening the fingers, calculated as the ratio of the point of the maximum velocity to the amplitude. It is also attributed to changes in the speed of motion. In other words, it is considered that the higher the degree of atrophy of the medial temporal region, the more the movement speed adjustment is affected; therefore, it is possible that the variability of fingers increases with atrophy in the VOI. This parameter is similar to the findings obtained in an aforementioned study [17]. The results of this study believe that this parameter can reflect the aggravation of cognitive function.

Currently, several studies have reported the relationship between the medial temporal cortex, including the hippocampus, and neuropsychological test results in patients with AD. There have been several reports on the relationship between the medial temporal cortex and the MMSE, which is known for general cognitive function tests, the Hasegawa Simple Intelligence Scale [26], and the Alzheimer’s Disease Rating Scale [27], but no reports have examined the relationship between VSRAD and MoCA-J. In this study, we found a correlation between the Z-score, indicating the degree of atrophy of the medial temporal region, which is the region of interest for AD, and the MoCA-J analysis. Other analysis indices have shown a significant correlation with the percentage of gray matter and white matter atrophy in the whole brain. In the medial temporal cortex, such as the entorhinal cortex, atrophy has been reported to appear in the early stages of AD and MCI [28-30]. Previous studies have also reported a correlation between MMSE, a general cognitive function test, and the Z-score [26]. In the present study, MoCA-J also showed a correlation with the Z-score, as well as MMSE. Other indices were correlated with the percentage of total brain atrophy areas in the gray and white matters. The overall gray matter and white matters of the brain are thought to decrease with age [31]. These indices reflect the proportion of atrophy of the entire brain in the aforementioned matters and may be correlated with MoCA-J, which is an overall functional rating. In VSRAD, there are other indices of analysis besides the Z-score, and we believe that it is also useful to observe these indices.

This study has some limitations. First, the sample size was small, and the study was conducted in a single center; therefore, the influence of selection bias cannot be denied. Therefore, further multicenter studies with more participants are needed. Second, this study did not examine patients with MCI at the pre-stage of AD. Many changes to the brain have been reported since the onset of MCI, such as atrophy of the entorhinal cortex [29, 30] and a reduction in gray matter and white mass throughout the brain [32]. Therefore, it is necessary to examine the relationship between the brain and hand function in the MCI stage in the future.

This study examined the relationship between VSRAD, cognitive function, and finger function in patients with AD. The results showed a significant relationship between the SD of distance rate of velocity peak in extending movements, a parameter of the finger-tapping motion, and the Z-score. We also found an association between neuropsychological tests and the overall degree of brain atrophy. The primary advantage of this study is that it suggests that the SD of distance rate of velocity peak in extending movements extracted from finger taps may be a useful parameter for the early detection of AD and diagnosis of its severity. We want to expand our analysis to include MCI and community-dwelling elderly individuals and examine whether finger-tapping exercise is a useful tool for early detection in the future.

## Data Availability

All relevant data are within the manuscript and its Supporting Information files.

## Acknowledgments

We would like to thank the participants for their contribution. We would also like to thank Editage (www.editage.com) for English language editing.

## Supporting information

### S1 Table. Results of finger tapping (dominant hand)

AD, Alzheimer’s disease; max, maximum; min: minimum; Ave, average; SD, standard deviation; and CV, coefficient of variation

### S2 Table. Results of finger tapping (non-dominant hand)

AD, Alzheimer’s disease; max, maximum; min: minimum; Ave, average; SD, standard deviation; and CV, coefficient of variation

### S3 Table. Results of finger tapping (in-phase)

AD, Alzheimer’s disease; max, maximum; min, minimum; Ave, average; SD, standard deviation; and CV, coefficient of variation

Upper values represent those for the in-phase left hand.

Lower values represent those for the in-phase right hand.

Numbers 41–44 are parameters for both-hand tasks only.

### S4 Table. Results of finger tapping (anti-phase)

AD, Alzheimer’s disease; max, maximum; min, minimum; Ave, average; SD, standard deviation; and CV, coefficient of variation

Upper values represent those for the anti-phase left hand.

Lower values represent those for the anti-phase right hand.

Numbers 41–44 are parameters for both-hand tasks only.

