## Supplementary material for "Relationship between finger movement characteristics and voxel-based specific regional analysis systems for Alzheimer’s disease": S1 Table

| No | Parameter | Mean ± SD | Correlation coefficient | p | No | Parameter | Mean ± SD | Correlation coefficient | p |
| --- | --- | --- | --- | --- | --- | --- | --- | --- | --- |
|  |  | AD group | (r) |  |  |  | AD group | (r) |  |
| 1 | Max distance amplitude (mm) | 86.1 ± 64.2 | -.088 | 0.486 | 21 | SD of distance rate of velocity peak in extending movements | 0.08 ± 0.02 | .07 | 0.543 |
| 2 | Total travelling distance (m) | 5231.2 ± 2329.8 | -.044 | 0.725 | 22 | SD of distance rate of velocity peak in flexing movement | 0.08 ± 0.01 | .053 | 0.677 |
| 3 | Ave of local max distance (mm) | 49.3 ± 23.9 | -.041 | 0.744 | 23 | Max of acceleration amplitude (m/s²) | 217.6 ± 364.3 | -.084 | 0.508 |
| 4 | SD of local max distance (mm) | 10.9 ± 11.2 | -.119 | 0.348 | 24 | Ave of local max acceleration in extending movement (m/s²) | 26.9 ± 13.3 | -.011 | 0.93 |
| 5 | Slope of approximate line of local max points (mm/s) | -0.03 ± 1.1 | .106 | 0.402 | 25 | Ave of local min acceleration in extending movement (m/s²) | −28.7 ± 15.7 | .086 | 0.499 |
| 6 | CV of local max distance | 0.2 ± 0.1 | -.08 | 0.528 | 26 | Ave of local max acceleration in flexing movement (m/s²) | 48.2 ± 26.5 | -.088 | 0.491 |
| 7 | SD of local max distance in three adjacent taps (mm) | 7.0 ± 7.1 | -.122 | 0.336 | 27 | Ave of local min acceleration in flexing movement (m/s²) | −39.9 ± 26.6 | .14 | 0.269 |
| 8 | Max of velocity amplitude (m/s) | 4.0 ± 5.4 | -.081 | 0.525 | 28 | Ave of contact duration (s) | 0.1 ± 0.02 | -.159 | 0.21 |
| 9 | Ave of local max velocity (m/s) | 0.8 ± 0.3 | -.094 | 0.458 | 29 | SD of contact duration (s) | 0.02 ± 0.01 | -.073 | 0.566 |
| 10 | Ave of local min velocity (m/s) | -1.1 ± 0.5 | .074 | 0.562 | 30 | CV of contact duration | 0.2 ± 0.06 | .042 | 0.742 |
| 11 | SD of local max velocity (m/s) | 0.2 ± 0.4 | -.119 | 0.349 | 31 | Number of zero crossover points of acceleration | 2.7 ± 0.8 | -.019 | 0.879 |
| 12 | SD of local min velocity (m/s) | 0.3 ± 0.5 | -.113 | 0.374 | 32 | Number of freezing calculated from acceleration | 18.7 ± 17.9 | .051 | 0.691 |
| 13 | Energy balance | 0.7 ± 0.1 | .049 | 0.702 | 33 | Number of taps | 52.6 ± 11.4 | .095 | 0.454 |
| 14 | Total energy (m²/s²) | 512.6 ± 612.7 | -.15 | 0.236 | 34 | Ave of tapping interval (s) | 0.2 ± 0.06 | -.16 | 0.207 |
| 15 | CV of local max velocity | 0.2 ± 0.2 | -.049 | 0.702 | 35 | Frequency of taps (Hz) | 3.5 ± 0.7 | .093 | 0.466 |
| 16 | CV of local min velocity | -0.2 ± 0.2 | .081 | 0.572 | 36 | SD of inter-tapping interval (s) | 0.05 ± 0.07 | -.126 | 0.319 |
| 17 | Number of freezing calculated from velocity | 7.1 ± 11.1 | .181 | 0.151 | 37 | CV of inter-tapping interval | 0.1 ± 0.2 | -.081 | 0.523 |
| 18 | Ave distance rate of velocity peak in extending movement | 0.5 ± 0.07 | -.067 | 0.601 | 38 | Inter-tapping interval variability (mm²) | 0.009 ± 0.05 | -.145 | 0.252 |
| 19 | Ave distance rate of velocity peak in flexing movement | 0.3 ± 0.06 | <.001 | 0.995 | 39 | Skewness of inter-tapping interval distribution | 1.1 ± 1.6 | .039 | 0.758 |
| 20 | Ratio of distance rates of velocity peak in extending and flexing movements | 1.3 ± 0.1 | -.08 | 0.53 | 40 | SD of inter-tapping interval in three adjacent taps (s) | 0.03 ± 0.03 | -.054 | 0.671 |
