## Supplementary material for "Relationship between finger movement characteristics and voxel-based specific regional analysis systems for Alzheimer’s disease": S2 Table

| No | Parameter | Mean ± SD | Correlation coefficient | p | No | Parameter | Mean ± SD | Correlation coefficient | p |
| --- | --- | --- | --- | --- | --- | --- | --- | --- | --- |
|  |  | AD group | (r) |  |  |  | AD group | (r) |  |
| 1 | Max distance amplitude (mm) | 79.5 ± 42.0 | -.030 | 0.817 | 21 | SD of distance rate of velocity peak in extending movements | 0.089 ± 0.021 | .515 | <.001 |
| 2 | Total travelling distance (m) | 4821.9 ± 2300.9 | .057 | 0.654 | 22 | SD of distance rate of velocity peak in flexing movement | 0.092 ± 0.022 | .054 | 0.67 |
| 3 | Ave of local max distance (mm) | 49.0 ± 25.4 | .095 | 0.453 | 23 | Max of acceleration amplitude (m/s²) | 128.0 ± 89.0 | -.174 | 0.169 |
| 4 | SD of local max distance (mm) | 9.7 ± 4.6 | .264 | 0.035 | 24 | Ave of local max acceleration in extending movement (m/s²) | 22.5 ± 8.9 | .054 | 0.672 |
| 5 | Slope of approximate line of local max points (mm/s) | -0.55 ± 1.23 | -.113 | 0.372 | 25 | Ave of local min acceleration in extending movement (m/s²) | −25.3 ± 11.8 | .041 | 0.748 |
| 6 | CV of local max distance | 0.22 ± 0.09 | .193 | 0.126 | 26 | Ave of local max acceleration in flexing movement (m/s²) | 41.6 ± 19.4 | -.012 | 0.925 |
| 7 | SD of local max distance in three adjacent taps (mm) | 6.6 ± 2.8 | .219 | 0.082 | 27 | Ave of local min acceleration in flexing movement (m/s²) | −32.9 ± 14.0 | .098 | 0.443 |
| 8 | Max of velocity amplitude (m/s) | 2.7 ± 1.5 | -.120 | 0.347 | 28 | Ave of contact duration (s) | 0.12 ± 0.03 | -.004 | 0.975 |
| 9 | Ave of local max velocity (m/s) | 0.77 ± 0.32 | .023 | 0.859 | 29 | SD of contact duration (s) | 0.036 ± 0.020 | .076 | 0.549 |
| 10 | Ave of local min velocity (m/s) | -1.00 ± 0.45 | -.003 | 0.979 | 30 | CV of contact duration | 0.27 ± 0.11 | .091 | 0.475 |
| 11 | SD of local max velocity (m/s) | 0.15 ± 0.06 | .143 | 0.26 | 31 | Number of zero crossover points of acceleration | 3.0 ± 1.5 | -.132 | 0.298 |
| 12 | SD of local min velocity (m/s) | 0.20 ± 0.08 | .215 | 0.088 | 32 | Number of freezing calculated from acceleration | 19.8 ± 17.5 | .029 | 0.822 |
| 13 | Energy balance | 0.78 ± 0.16 | -.096 | 0.452 | 33 | Number of taps | 48.5 ± 11.7 | -.004 | 0.978 |
| 14 | Total energy (m²/s²) | 387.0 ± 354.8 | .021 | 0.87 | 34 | Ave of tapping interval (s) | 0.32 ± 0.09 | -.129 | 0.31 |
| 15 | CV of local max velocity | 0.21 ± 0.06 | .125 | 0.326 | 35 | Frequency of taps (Hz) | 3.2 ± 0.77 | -.003 | 0.98 |
| 16 | CV of local min velocity | -0.21 ± 0.07 | -.173 | 0.171 | 36 | SD of inter-tapping interval (s) | 0.07 ± 0.17 | -.164 | 0.194 |
| 17 | Number of freezing calculated from velocity | 7.4 ± 10.4 | .047 | 0.712 | 37 | CV of inter-tapping interval | 0.20 ± 0.20 | -.072 | 0.57 |
| 18 | Ave distance rate of velocity peak in extending movement | 0.50 ± 0.06 | -.050 | 0.694 | 38 | Inter-tapping interval variability (mm²) | 0.008 ± 0.032 | -.107 | 0.402 |
| 19 | Ave distance rate of velocity peak in flexing movement | 0.39 ± 0.06 | -.051 | 0.689 | 39 | Skewness of inter-tapping interval distribution | 1.45 ± 1.60 | .136 | 0.283 |
| 20 | Ratio of distance rates of velocity peak in extending and flexing movements | 1.3 ± 0.2 | .008 | 0.947 | 40 | SD of inter-tapping interval in three adjacent taps (s) | 0.04 ± 0.06 | -.090 | 0.479 |
