## Supplementary material for "Relationship between finger movement characteristics and voxel-based specific regional analysis systems for Alzheimer’s disease": S3 Table

| No | Parameter | Mean ± SD AD group | Correlation  coefficient (r) | p | No | Parameter | Mean ± SD AD group | Correlation  coefficient (r) | p |
| --- | --- | --- | --- | --- | --- | --- | --- | --- | --- |
| 1 | Max distance amplitude (mm) | 72.3 ± 28.4 79.4 ± 56.7 | .052 .032 | 0.684 0.803 | 23 | Max acceleration amplitude (m/s²) | 125.5 ± 57.4 200.4 ± 359.2 | -.042 -.007 | 0.74 0.957 |
| 2 | Total travelling distance (m) | 4878.9 ± 1992.0 4900.7 ± 2092.4 | .068 -.007 | 0.595 0.958 | 24 | Ave of local max acceleration in extending movement (m/s²) | 22.5 ± 8.2 23.1 ± 12.0 | .11 <.001 | 0.384 1 |
| 3 | Ave of local max distance (mm) | 48.1 ± 20.9 47.4 ± 22.6 | .044 -.002 | 0.729 0.988 | 25 | Ave of local min acceleration in extending movement (m/s²) | −26.1 ± 11.8 −26.0 ± 13.5 | -.044 .094 | 0.727 0.457 |
| 4 | SD of local max distance (mm) | 8.6 ± 3.8 9.9 ± 10.1 | .127 .014 | 0.315 0.916 | 26 | Ave of local max acceleration in flexing movement (m/s²) | 43.3 ± 17.1 45.05 ± 25.8 | .058 .019 | 0.649 0.879 |
| 5 | Slope of approximate line of local max points (mm/s) | −0.4 ± 0.8 ∍−0.4 ± 1.4 | -.064 -.11 | 0.614 0.931 | 27 | Ave of local min acceleration in flexing movement (m/s²) | −34.3 ± 13.1 ∍−37.9 ± 23.7 | .02 .029 | 0.875 0.818 |
| 6 | CV of local max distance | 0.1 ± 0.06 0.2 ± 0.1 | .08 .054 | 0.527 0.669 | 28 | Ave of contact duration (s) | 0.1 ± 0.03 0.1 ± 0.02 | -.081 -.089 | 0.523 0.485 |
| 7 | SD of local max distance in three adjacent taps (mm) | 5.8 ± 2.6 6.2 ± 5.0 | .058 .022 | 0.647 0.865 | 29 | SD of contact duration (s) | 0.03 ± 0.02 0.02 ± 0.02 | -.071 -.048 | 0.578 0.709 |
| 8 | Max velocity amplitude (m/s) | 2.6 ± 1.0 3.5 ± 4.8 | .014 .012 | 0.915 0.928 | 30 | CV of contact duration | 0.2 ± 0.09 0.2 ± 0.1 | -.07 -.032 | 0.58 0.80 |
| 9 | Ave of local max velocity (m/s) | 0.7 ± 0.2 0.7 ± 0.3 | .022 -.028 | 0.864 0.824 | 31 | Number of zero crossover points of acceleration | 2.7 ± 0.9 2.6 ± 0.6 | -.001 .063 | 0.99 0.62 |
| 10 | Ave of local min velocity (m/s) | −1.0 ± 0.3 −1.0 ± 0.5 | -.052 -.015 | 0.681 0.907 | 32 | Number of freezing calculated from acceleration | 17.2 ± 16.7 15.8 ± 14.9 | .075 .103 | 0.553 0.418 |
| 11 | SD of local max velocity (m/s) | 0.1 ± 0.06 0.2 ± 0.4 | -.02 -.021 | 0.877 0.869 | 33 | Number of taps | 49.9 ± 9.8 50.6 ± 9.4 | .035 .071 | 0.783 0.577 |
| 12 | SD of local min velocity (m/s) | 0.1 ± 0.08 0.2 ± 0.5 | .163 .008 | 0.197 0.948 | 34 | Ave of tapping interval (s) | 0.3 ± 0.06 0.3 ± 0.05 | -.085 -.102 | 0.502 0.423 |
| 13 | Energy balance | 0.7 ± 0.1 0.7 ± 0.1 | -.031 -.03 | 0.81 0.813 | 35 | Frequency of taps (Hz) | 3.3 ± 0.6 3.4 ± 0.6 | .03 .063 | 0.811 0.621 |
| 14 | Total energy (m²/s²) | 378.2 ± 286.9 443.4 ± 533.7 | .052 .017 | 0.686 0.892 | 36 | SD of inter-tapping interval (s) | 0.04 ± 0.04 0.03 ± 0.02 | .011 .012 | 0.934 0.922 |
| 15 | CV of local max velocity | 0.1 ± 0.05 0.2 ± 0.2 | -.07 -.016 | 0.583 0.9 | 37 | CV of inter-tapping interval | 0.1 ± 0.09 0.1 ± 0.07 | .058 .024 | 0.649 0.854 |
| 16 | CV of local min velocity | −0.1 ± 0.06 −0.2 ± 0.2 | -.109 .025 | 0.389 0.846 | 38 | Inter-tapping interval variability (mm²) | 0.004 ± 0.02 0.001 ± 0.002 | -.024 .055 | 0.848 0.665 |
| 17 | Number of freezing calculated from velocity | 5.3 ± 8.4 5.7 ± 8.2 | .16 .163 | 0.206 0.197 | 39 | Skewness of inter-tapping interval distribution | 0.9 ± 1.4 0.8 ± 1.3 | .085 -.122 | 0.505 0.335 |
| 18 | Ave distance rate of velocity peak in extending movement | 0.5 ± 0.06 0.5 ± 0.08 | .086 -.037 | 0.498 0.774 | 40 | SD of inter-tapping interval in three adjacent taps (s) | 0.03 ± 0.03 0.02 ± 0.01 | .002 .044 | 0.985 0.727 |
| 19 | Ave distance rate of velocity peak in flexing movement | 0.3 ± 0.06 0.3 ± 0.07 | -.125 -.062 | 0.324 0.627 | 41 | Ave phase difference between the left hand and right-hand tapping (degree) | −10.5 ± 30.7 | -.234 | 0.062 |
| 20 | Ratio of distance rates of velocity peak in extending and flexing movements | 1.3 ± 0.2 1.3 ± 0.2 | 0.164 0.025 | 0.195 0.842 | 42 | SD of phase difference between the left hand and right-hand tapping (degree) | 43.4 ± 24.9 | .039 | 0.758 |
| 21 | SD of distance rate of velocity peak in extending movements | 0.09 ± 0.02 0.08 ± 0.03 | 0.515 0.077 | <0.001 0.543 | 43 | Similarity of hands | 0.7 ± 0.2 | -.015 | 0.908 |
| 22 | SD of distance rate of velocity peak in flexing movement | 0.08 ± 0.01 0.08 ± 0.02 | .011 -.039 | 0.934 0.759 | 44 | Time lag of similarity of hands (s) | 0.007 ± 0.02 | .199 | 0.114 |
