## Supplementary material for "Relationship between finger movement characteristics and voxel-based specific regional analysis systems for Alzheimer’s disease": S4 Table

| No | Parameter | Mean ± SD AD group | Correlation  coefficient (r) | p | No | Parameter | Mean ± SD AD group | Correlation  coefficient (r) | p |
| --- | --- | --- | --- | --- | --- | --- | --- | --- | --- |
| 1 | Max distance amplitude (mm) | 111.1 ± 52.4 125.0 ± 69.8 | .131 .131 | 0.299 0.299 | 23 | Max of acceleration amplitude (m/s²) | 181.5 ± 254.1 259.5 ± 386.3 | .078 .166 | 0.536 0.189 |
| 2 | Total travelling distance (m) | 3816.7 ± 1790.4 4076.4 ± 2166.2 | .063 .036 | 0.618 0.775 | 24 | Ave of local max acceleration in extending movement (m/s²) | 22.4 ± 10.2 27.8 ± 29.6 | .183 .158 | 0.145 0.210 |
| 3 | Ave of local max distance (mm) | 77.1 ± 26.4 82.2 ± 35.7 | .130 .091 | 0.303 0.470 | 25 | Ave of local min acceleration in extending movement (m/s²) | −15.7 ± 8.0 −15.0 ± 12.9 | -.057 -.090 | 0.65 0.474 |
| 4 | SD of local max distance (mm) | 12.9 ± 9.4 17.2 ± 18.6 | .168 .192 | 0.181 0.126 | 26 | Ave of local max acceleration in flexing movement (m/s²) | 50.6 ± 18.3 61.2 ± 34.3 | .129 .120 | 0.305 0.341 |
| 5 | Slope of approximate line of local max points (mm/s) | -0.38 ± 1.28 -0.58 ± 1.96 | -.0450 -.0550 | 0.719 0.664 | 27 | Ave of local min acceleration in flexing movement (m/s²) | −28.3 ± 12.4 −39.8 ± 34.7 | -.062 -.198 | 0.624 0.115 |
| 6 | CV of local max distance | 0.18 ± 0.14 0.21 ± 0.17 | .097 .117 | 0.442 0.354 | 28 | Ave of contact duration (s) | 0.34 ± 0.26 0.33 ± 0.32 | .130 .120 | 0.302 0.340 |
| 7 | SD of local max distance in three adjacent taps (mm) | 8.9 ± 5.9 12.8 ± 15.5 | .163 .206 | 0.196 0.101 | 29 | SD of contact duration (s) | 0.141 ± 0.284 0.125 ± 0.204 | .251 .239 | 0.044 0.056 |
| 8 | Max of velocity amplitude (m/s) | 3.9 ± 4.7 5.0 ± 6.5 | .089 .175 | 0.482 0.164 | 30 | CV of contact duration | 0.31 ± 0.21 0.32 ± 0.23 | .180 ,129 | 0.153 0.307 |
| 9 | Ave of local max velocity (m/s) | 0.81 ± 0.29 0.91 ± 0.55 | .161 .208 | 0.201 0.098 | 31 | Number of zero crossover points of acceleration | 11.4 ± 14.4 10.9 ± 12.6 | .047 .038 | 0.706 0.763 |
| 10 | Ave of local min velocity (m/s) | -1.25 ± 0.42 -1.50 ± 0.83 | -.140 -.137 | 0.269 0.277 | 32 | Number of freezing calculated from acceleration | 62.3 ± 38.4 62.6 ± 37.1 | .123 .053 | 0.330 0.676 |
| 11 | SD of local max velocity (m/s) | 0.26 ± 0.48 0.42 ± 0.92 | .112 .223 | 0.375 0.074 | 33 | Number of taps | 24.1 ± 10.9 24.2 ± 10.6 | -.025 -.008 | 0.841 0.944 |
| 12 | SD of local min velocity (m/s) | 0.33 ± 0.40 0.56 ± 1.03 | .120 .244 | 0.343 0.050 | 34 | Ave of tapping interval (s) | 0.78 ± 0.54 0.73 ± 0.40 | .104 .035 | 0.409 0.782 |
| 13 | Energy balance | 0.68 ± 0.18 0.63 ± 0.32 | .105 -.067 | 0.408 0.595 | 35 | Frequency of taps (Hz) | 1.5 ± 0.72 1.6 ± 0.70 | -.044 -.013 | 0.728 0.913 |
| 14 | Total energy (m²/s²) | 332.9 ± 262.9 452.0 ± 558.4 | .101 .128 | 0.423 0.310 | 36 | SD of inter-tapping interval (s) | 0.19 ± 0.28 0.18 ± 0.25 | .225 .034 | 0.073 0.783 |
| 15 | CV of local max velocity | 0.29 ± 0.43 0.31 ± 0.35 | .102 .157 | 0.421 0.214 | 37 | CV of inter-tapping interval | 0.22 ±0.13  0.21 ± 0,17 | .237 .077 | 0.058 0.542 |
| 16 | CV of local min velocity | -0.27 ± 0.26 -0.29 ± 0.31 | -.095 -.231 | 0.452 0.064 | 38 | Inter-tapping interval variability (mm²) | 0.021 ± 0.040 0.038 ± 1.137 | .056 .052 | 0.654 0.680 |
| 17 | Number of freezing calculated from velocity | 17.0 ± 14.7 15.0 ± 12.2 | .087 .011 | 0.492 0.930 | 39 | Skewness of inter-tapping interval distribution | 0.74 ± 0.96 0.65 ± 1.10 | .082 -.123 | 0.517 0.332 |
| 18 | Ave distance rate of velocity peak in extending movement | 0.41 ± 0.07 0.40 ± 0.07 | .109 .082 | 0.391 0.518 | 40 | SD of inter-tapping interval in three adjacent taps (s) | 0.16 ± 0.29 0.14 ± 0.23 | .209 .070 | 0.095 0.580 |
| 19 | Ave distance rate of velocity peak in flexing movement | 0.34 ± 0.06 0.35 ± 0.07 | -.240 .064 | 0.055 0.611 | 41 | Ave phase difference between the left hand and right-hand tapping (degree) | 183.7 ± 54.3 | -.064 | 0.61 |
| 20 | Ratio of distance rates of velocity peak in extending and flexing movements | 1.2 ± 0.3 1.1 ± 0.2 | .240 .097 | 0.055 0.441 | 42 | SD of phase difference between the left hand and right-hand tapping (degree) | 57.8 ± 47.9 | -.013 | 0.912 |
| 21 | SD of distance rate of velocity peak in extending movements | 0.094 ± 0.036 0.100 ± 0.048 | .034 .140 | 0.789 0.268 | 43 | Similarity of hands | 0.21 ± 0.25 | -.048 | 0.701 |
| 22 | SD of distance rate of velocity peak in flexing movement | 0.094 ± 0.034 0.097 ± 0.039 | -.077 -.049 | 0.541 0.697 | 44 | Time lag of similarity of hands (s) | 0.042 ± 0.327 | .001 | 0.99 |
